# CX3CR1 as a Respiratory Syncytial Virus Receptor in Pediatric Human Lung

**DOI:** 10.1101/19002394

**Authors:** Christopher S. Anderson, Chin-Yi Chu, Qian Wang, Jared A. Mereness, Yue Ren, Kathy Donlon, Soumyaroop Bhattacharya, Ravi S. Misra, Edward E. Walsh, Gloria S. Pryhuber, Thomas J. Mariani

**Author notes:** Corresponding Author Address for Correspondence: Thomas J Mariani, PhD, Division of Neonatology and Pediatric Molecular and Personalized Medicine Program, University of Rochester Medical Center, 601 Elmwood Ave, Box 850, Rochester, NY 14642, USA. None of the authors have any conflicts of interest, financial or otherwise, to declare. **Author Contributions** Each author has met the Pediatric Research authorship requirements. CSA, EEW, and TJM conceptualized the study. CSA, TJM, CC, QW, EEW, JAM, YR, and GP designed the experiments. TJM, GP, and RM developed the cohort, and collected the specimens. CSA, KD, TJM, CC, JM, YR, and SB generated, analyzed and interpreted the data. CSA, CC, JAM, GP EEW and TJM wrote and/or revised the manuscript. - We showed that RSV demonstrated preferential infection of CX3CR1 positive pediatric epithelial cells - Blocking CX3CR1/RSV interaction significantly decreased productive viral infection in vitro - We found that CX3CR1 transcript are often expressed in both the upper (60%) and lower airways (36%) of pediatric subjects - CX3CR1 tissue localization and intracellular transcript in donor quality pediatric lung specimens, demonstrating an apical RSV receptor on pediatric airways for the first time. - These data demonstrate CX3CR1 is both present in the airways of pediatric subjects where it may serve as a receptor for RSV infection, and plays a mechanistic role in mediating viral infection of pediatric airway epithelial cells in vitro.

## Abstract

**BACKGROUND:** Data on the host factors that contribute to infection of young children by Respiratory Syncytial Virus (RSV) are limited. The human chemokine receptor, CX3CR1, has recently been implicated as an RSV receptor. Here we evaluate a role for CX3CR1 in pediatric lung RSV infections.

**METHODS:** CX3CR1 transcript levels in the upper and lower pediatric airways were assessed. Tissue localization and cell specific expression was confirmed using *in situ* hybridization and immunohistochemistry. The role of CX3CR1 in RSV infection was also investigated using a novel physiological model of pediatric epithelial cells.

**RESULTS:** Low levels of CX3CR1 transcript were often, but not always, expressed in both upper (62%) and lower airways (36%) of pediatric subjects. CX3CR1 transcript and protein expression was detected in epithelial cells of normal human pediatric lung tissues. CX3CR1 expression was readily detected on primary cultures of differentiated pediatric/infant human lung epithelial cells. RSV demonstrated preferential infection of CX3CR1 positive cells, and blocking CX3CR1/RSV interaction significantly decreased viral load.

**CONCLUSION:** CX3CR1 is present in the airways of pediatric subjects where it may serve as a receptor for RSV infection. Furthermore, CX3CR1 appears to play a mechanistic role in mediating viral infection of pediatric airway epithelial cells in vitro.

## Introduction

Almost all children are infected with Respiratory Syncytial Virus (RSV) during their first two years of life. RSV infection typically originates in the upper airways of young children and can be found in the lower airways during severe cases. RSV replication in the airways occurs primarily in epithelial cells and these cells are readily infected *in vitro*. The innate host properties that make epithelial cells prone to RSV infection are still not well understood^1,2^.

To date, there is no consensus on a specific host receptor that RSV uses for attachment to the host epithelium to initiate infection. Studies of RSV attachment have demonstrated that addition of heparin, heparan sulfates, or chondroitin to submerged cell lines significantly decreases RSV-cell association^3^. Other studies have shown that this interaction was specific to particular heparan sulfate and chondroitin molecules ^1^. These findings have been complicated by contradictory results of studies of heparan sulfate proteoglycans (HSPG) expression on the lung epithelium, with some reporting the lack of expression and others noting expression on normal human bronchial epithelial cells grown *in vitro* ^4^.

The fractalkine receptor, CX3CR1, is a 7-transmembrane G protein-coupled receptor (GPCR) known to be expressed in NK cells, cytotoxic CD8 T cells, monocytes, and dendritic cells. The CX3CR1 ligand, CX3CL1 (fractalkine), contains a CX3C motif and can be expressed on the cellular membrane or as a soluble form. Fractalkine is expressed in the lung, and is thought to play a role in migration and retention of immune cells in the tissue ^5^. The presence of a CX3C motif and mucin-like regions of the RSV G-protein has led to interest in the involvement of CX3CR1 in RSV infection. RSV infection of *in vitro* models of adult epithelium have shown at least partial dependence upon CX3CR1/G-protein binding ^6,7^. RSV replication has been shown to occur in ciliated cells in animal models ^8^, and CX3CR1 and cilia have been shown to co-localize in adult epithelial cells *in vitro* ^9^. RSV strains containing a CX3C to CX4C motif mutation show reduced replication *in vitro*^6^. Furthermore, prophylactic treatment with monoclonal antibodies targeting the CX3C motif has been shown to reduce RSV disease in mice ^10,11^. Taken together, the CX3C motif of the RSV G-protein is emerging as a CX3CR1 ligand that impacts infection by RSV.

Despite the clinical significance of RSV infection in children, no definitive studies have assessed the importance of CX3CR1 in pediatric RSV infection. Moreover, the extent of CX3CR1 expression in pediatric airways in humans is unclear. Using our unique access to normal newborn and pediatric human lung tissues, as well as our newly developed pediatric lung epithelial cell model, we set out to examine the expression of CX3CR1 in the pediatric lung and test its role in RSV infection of pediatric airways.

## Methods

### Virus Propagation

The GFP containing RSV (A2 strain) ^12^ was grown in Hep-2 cells as previously described^13^. Cells were incubation at 37°C for 2hrs with, supernatant was removed, and 5ml of virus medium was added to the flask. Virus was allowed to propagate for 5-7 days before cytopathic effect was observed. Supernatant was aspirated, centrifuged at 300g for 10 minutes to remove cell debris. Cleared supernatant was aliquoted into cryopreserved vials. Vials were immediately flash frozen in liquid nitrogen and stored at -80°C until usage.

### Plaque Forming Units (PFU) Quantification

To quantify virus plaque forming units (PFU). Hep-2 cells were seeded onto 96-well plates (Costar 3596) at a density of 2.5 × 10^4^ cells per well in 200uL of Hep-2 media (2.5×10^6^ per 100 wells in 20ml). The next day, 0.6% Agarose (Sigma) in molecular grade H20 was heated in a microwave until melted (approximately 2 minutes) and place in 42°C water bath. Next, virus containing supernatant was quickly thawed at 37°C in a water bath and a ten-fold dilution series was performed resulting in 11 dilutions. Cells were subsequently washed with DPBS(+Ca+Mg). Virus (100ul at each dilution) was added to wells in duplicate. Additionally, 100ul of virus media was used as a negative control. Inoculated cells were incubated at 37°C for 1hr and rocked every 15 minutes to keep the culture covered and wet. After the 1hr incubation, virus supernatant was carefully removed. Heated (42°C) agarose was diluted with Hep-2 growth medium at a ratio of 1:1 and quickly overlaid on top of the cells. Cultures were maintained at room temperature for ten minutes to allow agarose to solidify. Cultures were incubated for 7 days at 37°C. Plaques were identified using fluorescent microscopy and counted to determine PFU/ml.

### Primary Pediatric Human Epithelial Cell Model

Pediatric human lung epithelial (PHLE) cells were prepared as previously described^14^. Briefly, fresh donor-quality infant/pediatric lung tissues were digested with a protease cocktail containing collagenase type A (2 mg/ml), dispase II (1 mg/ml), elastase (0.5 mg/ml) and DNAase (2 mg/ml) ^15^. Single cell suspensions were transferred to a T75 flask containing SAGM (Lonza) supplemented with 1% FBS. After 24 hours, non-adherent cells were removed. Fibroblast-like cells were removed periodically by treatment with 0.0125% trypsin containing 45μM EDTA in DPBS at room temperature. Once cells were 60-70% confluent, cells were trypsinized and 100,000 cells were seeded on rat tail collagen-I (34.5 μg/mL) coated Transwell inserts (12 well PET membrane, 0.4 μm pore size, 12 mm diameter). After 24 hours, culture medium was changed to 1:1 mixture of BEBM(Lonza)/DMEM. After confluence (10-15 days), resistance was measured daily until resistance reached ≥ 200 Ohms*cm^2^, the apical medium was removed and cells were maintained at air-liquid interface (ALI). After transition to air-liquid interface, cells were maintained in PneumaCult-ALI medium containing supplements and hydrocortisone, according to manufacturer’s instructions. ALI cultures were differentiated for 12-14 days at ALI.

### Virus Infection

Differentiated (12-14 day ALI) PHLE cells were infected (MOI of 0.5-1) were added to the apical surface, and incubated for 2 hours at 37°C. Virus were removed, and cells were washed with PBS, and returned to the incubator for up to 48 hrs.

### Immunohistochemistry

Formalin fixed paraffin embedded (FFPE) lung tissues were obtained from the Human Tissue Core of the Developing Lung Molecular Atlas Program (LungMAP HTC). Tissue was deparaffinization by washing slides sequentially twice in Xylene, a 1:1 mixture of 100% ethanol/Xylene once, twice with 100% ethanol, once with 95% ethanol, once with 70% ethanol, once with 50% ethanol, each for 3 minutes, and finally rinsed with cold tap water. For antigen retrieval, slides were placed in an open vessel inside a vegetable steamer and covered with boiling sodium citrate buffer (10 mM sodium citrate, 0.05% Tween 20, pH 6.0) for 20 minutes. Slides were rinsed with running cold tap water for 10 min and washed twice for 5 min in Tris-buffered saline (TBS) containing 0.025% Triton X-100 with gentle agitation, and subsequently blocked in 10% normal goat serum with 1% BSA in TBS for 2 h at room temperature. Slides were drained and excess liquid removed. Rabbit anti-CX3CR1 antibody (Abcam ab8021) was diluted 1:50 in TBS with 1% BSA, and mixed with mouse anti-acetylated tubulin diluted 1:100. 200ul of diluted antibody mixture was added to each slide and incubated overnight at 4°C. Normal mouse IgG and goat IgG served as negative controls. After overnight incubation, slides were rinsed twice for 5 min with TBS with 0.025% Triton, with gentle agitation. Fluorophore-conjugated anti-mouse and anti-rabbit secondary antibody was diluted 1:1000 in TBS with 1% BSA. 200ul of diluted secondary antibody mixture was added to each slide, and incubated for 1 h at room temperature in the dark. After incubation, slides were rinse three times for 5 min with TBS. Slides were mounted using ProLong Gold Antifade mounting medium (Life Technologies). Slides were incubated overnight at RT to allow slides to cure.

### Fluorescent *in Situ* Hybridization

Fluorescent *in Situ* Hybridization (FISH) was performed using RNAscope technology (ACDbio), essentially according to the manufacturer’s instructions. In brief, FFPE lung tissues were deparaffinized, target retrieved, and probed with CX3CR1 (ACDbio cat# 411251), FOXJ1 (ACDbio cat# 430921-C2), or SCGB1A1 (ACDbio cat# 469971-C3) transcript specific probes using the RNAscope Multiplex Fluorescent Kit v2 (ACDbio cat# 323100) and Opal Fluorophores (PerkinElmer Opal 520, Opal 570, Opal 690, respectively).

### RNA Quantification

Pre-infection PHLE cells derived from four subjects were used for CX3CR1 RNA quantification and post infection PHLE cells were used for RSV-M protein quantification. PHLE cells were washed twice with PBS. Cells were lysed with 150ul of RNA lysis buffer(Agilent) and RNA was extracted using the Absolutely RNA Micoprep Kit (Agilent) according to the manufacture’s protocol, including DNase treatment. RNA was quantified by Nanodrop spectrometry and 500ug of RNA was used for cDNA synthesis (iScript; BIO-RAD). cDNA was diluted 1:20 with molecular grade H2O and 6ul of cDNA was added to 4ul Power SYBR green PCR Master Mix (Applied Biosystems) containing Human CX3CR1 (FWD: AGTGTCACCGACATTTACCTCC, REV: AAGGCGGTAGTGAATTTGCAC) or RSM M protein (FWD: GGCAAATATGGAAACATAGCTGAA, REV: TCTTTTTCTAGGACATTGTATTGA ACAG) primers. Quantitative PCR was performed using Viia7 (applied biosystems).

### CX3CR1 Protein Quantification

Pre-infection differentiated PHLE cells were washed with PBS and lysed with 50ul RIPA buffer containing protease inhibitors (Thermo Scientific) for 5 minutes on ice. Lysates were spun at 10,000g for 10 minutes and supernatant was collected. CX3CR1 protein levels were determine by Sandwich-ELISA (myBioSource, MBS2505819) following the manufacturer’s instructions.

### Flow Cytometry

Following RSV infection, cells were recovered by trypsinization (0.25%), blocked with BSA, and incubated with anti-CX3CR1 antibody (Abcam ab8021) for 1 hr at RT. Cells were analyzed for GFP and CX3CR1 on a BD LSRII. Data was analyzed using FlowJo Software.

### Statistics

Statistics were performed using the R software base package. Significance was achieved using a p value of 0.05 threshold.

## Results

We interrogated CX3CR1 expression in an RNAseq database we have recently generated, describing upper airway gene expression in nasal swabs obtained from infants at ∼1 month of age^16^. CX3CR1 transcript levels were detected in 33 out of 53 (62%) infant upper airway samples. CX3CR1 transcript levels varied between subjects ranging from 0.29 to 22.3 rpm-normalized counts in subjects with detected CX3CR1 transcript (Figure 1A). Among the 23,132 gene transcripts detected in any subject, mean CX3CR1 transcript level was greater than 44% of the expressed genes. Among chemokine receptors, CX3CR1 transcript ranked 9 out of 19 (47%; Figure 1B). Taken together, CX3CR1 transcript was detected in the majority of subjects, but not all, and the absolute expression levels varied widely.

**Figure 1.**
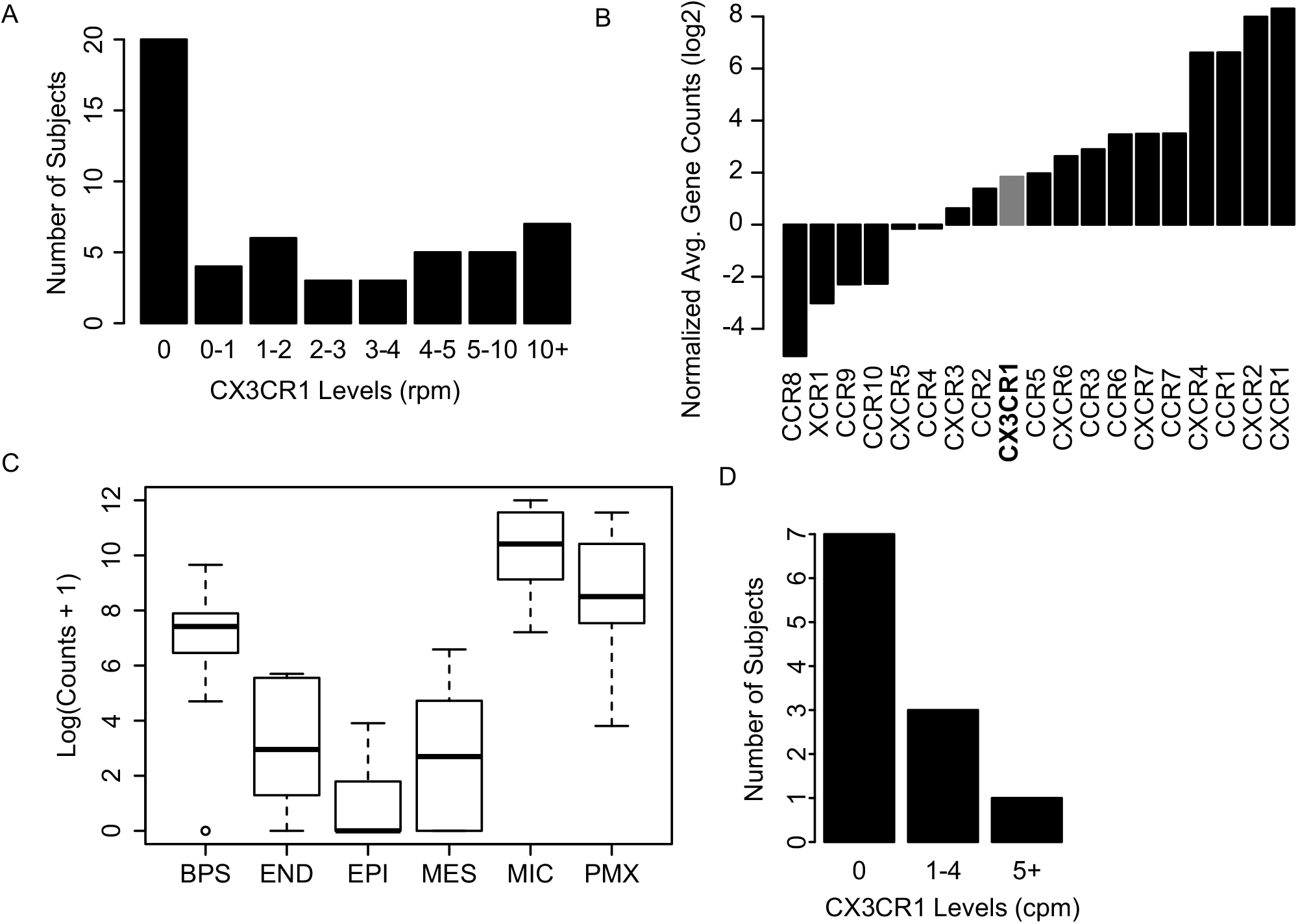
Pediatric Airway CX3CR1 Transcript Levels. (A) CX3CR1 transcript levels from RNA extracted from nasal swabs of young children. (B) Chemokine receptor transcript levels RNA extracted from nasal swabs of young children. (C) CX3CR1 gene expression from cells sorted after lung digest (BPS: biopsy, END: endothelial, EPI: epithelial, MES: mesenchymal, MIC: mixed immune cells, PMX: mixed pulmonary). (D) CX3CR1 transcript levels of sorted epithelial cells population.

We also interrogated CX3CR1 expression in a second database describing lower airway gene expression from 5 cell populations obtained from total tissue digests of infant/pediatric human lung lobes from LungMAP^15^; sorted epithelial (EPI; EPCAM+/CD45-), sorted endothelial (END; CD45-/CD31+), sorted mesenchymal cells (MES; CD45-/EPCAM-/CD31-), sorted mixed immune cells (MIC; CD45+), unsorted mixed pulmonary (PMX; unsorted), as well as tissues biopsies (BPS). CX3CR1 transcript was detected in all sample types (Figure 1C). The highest CX3CR1 expression was found in mixed immune cells, followed by biopsy and mesenchymal. CX3CR1 transcript in sorted epithelial cells was detected in 4 out of 11 (36%) subjects and when present, transcript counts were relatively low (1-6 CPM normalized counts). Taken together, CX3CR1 transcript was sometimes detected in lower respiratory track epithelial cells from pediatric lungs.

CX3CR1 expression in pediatric lungs was also assessed in fixed lung tissues. In situ hybridization of histological sections from normal pediatric lungs detected CX3CR1 transcript in epithelial cells lining the airway, including those expressing FOXJ1, a ciliated cell differentiation maker (Figure 2A). Immunohistochemistry for CX3CR1 protein also detected expression in airway epithelial cells (Figure 2B), where CX3CR1 co-localized with acetylated tubulin, consistent with *in vitro* studies in adults^9^. Taken together, CX3CR1 transcript and protein is detectable in epithelial cells lining the airway of intact pediatric lung tissues.

**Figure 2.**
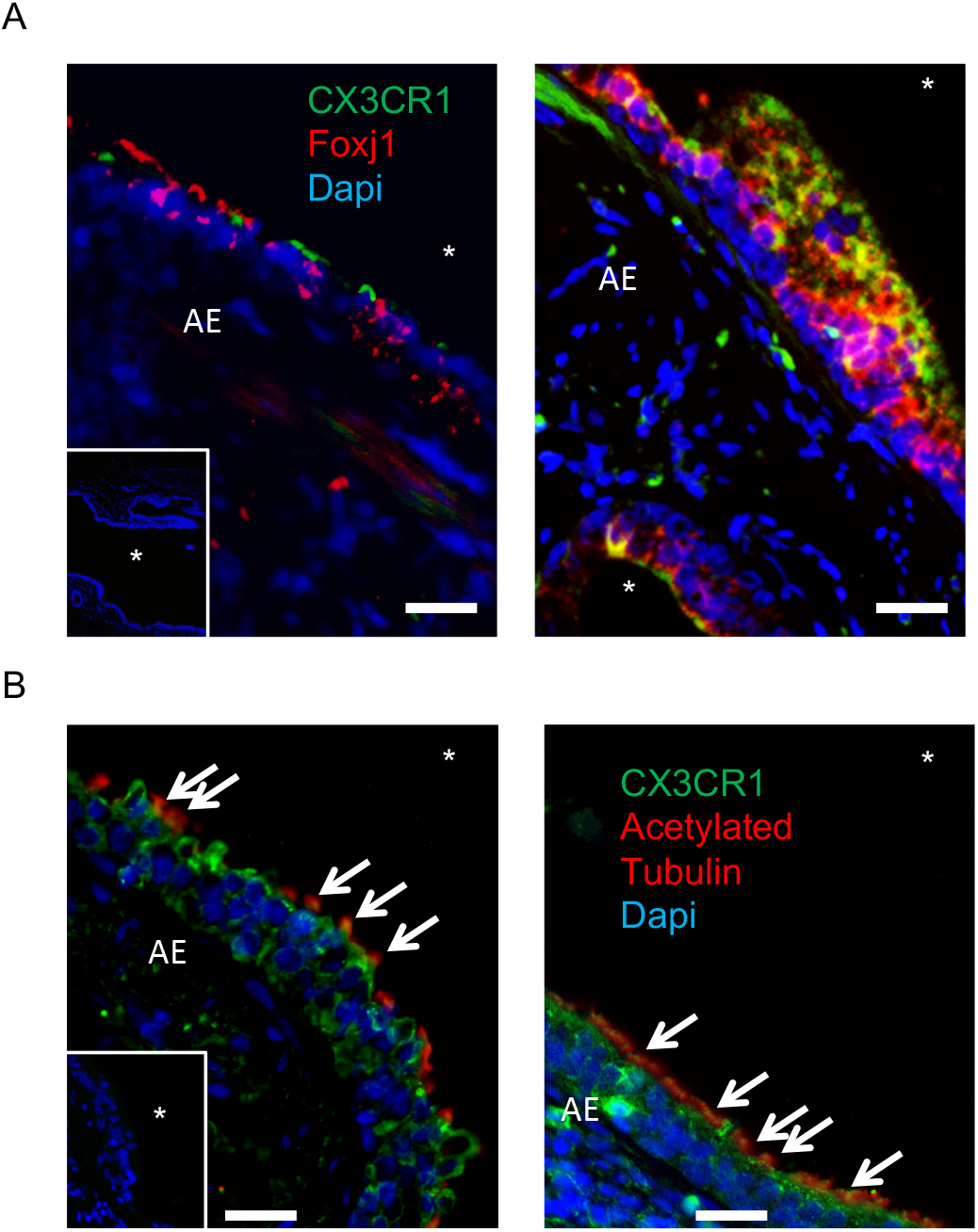
*In Situ* CX3CR1 Expression in the Pediatric Airways. (A) Fluorescent *in situ* hybridization of pediatric lung samples probed for CX3CR1 (green), Foxj1 (red), and nuclei (blue) for two subjects. In-laid image: negative probe straining for green and red probes and stained for nuclei (blue). (B) Lung specimens probed with anti-CX3CR1 antibody (green), anti-acetylated tubulin antibody (red), and counterstained with dapi nuclear stain (blue). In-laid image: negative control probed with unimmunized rabbit IgG (green) or mouse IgG (blue) and counterstained with nuclear stain (blue).

Next, we assessed the expression of CX3CR1 in primary cultures of pediatric human lung epithelial cells (PHLE). PHLE cells were differentiated at air-liquid interface *in vitro*, as previously described^14^, and compared to Hep-2 cells grown under standard submerged conditions. We found that CX3CR1 mRNA was consistently detected in both Hep-2 and PHLE cells, with PHLE cells expressing nearly 5-fold greater levels (Figure 3A). CX3CR1 transcript levels varied somewhat among PHLE cells derived from lung tissue of different donor organs, but this variation was similar to Hep-2 cell replicates. CX3CR1 protein levels were also readily detected in PHLE cultures from multiple donors, and were nearly 2-fold higher in PHLE cells compared to Hep-2 cells (Figure 3B). Taken together, CX3CR1 transcript and protein was detected in air-liquid interface cultures of differentiated pediatric lung epithelial cells.

**Figure 3.**
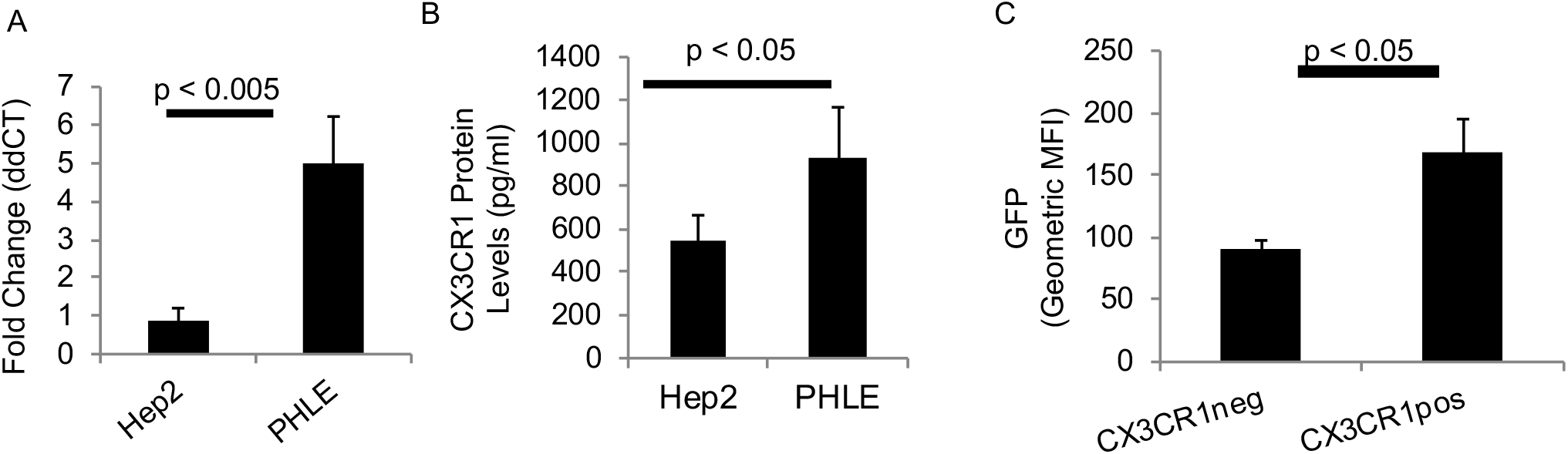
Role of CX3CR1 in RSV Infection of Pediatric Epithelial Cells. (A) RNA transcript levels of PHLE cells 14 days post ALI (N=4) measured by quantitative PCR for the CX3CR1 transcript. Hep-2 cell line (n=3) was used for comparison. (B) Protein levels of PHLE cells 14 days post ALI (N=4) measured by sandwich ELISA from cell protein lysates. Hep-2 cell line (n=3) was used for comparison. (C) Flow cytometric analysis of CX3CR1 positive and negative cells after infection of PHLE cells for 48hrs and with GFP expressing RSV (n=3).

We evaluated the functional role of CX3CR1 in RSV infection of pediatric lung epithelial cells. PHLE cells were differentiated at ALI as described, and challenged at the apical surface with RSV engineered to express GFP^17^. PHLE cells were readily infected, as defined by detection of fluorescence in less than 24 hrs, and lasting for more than 72 hrs, following challenge (Figure 4A, data not shown). Evidence of productive RSV infection in PHLE cells is demonstrated by detection of fluorescent foci and viral transcript (RSV-M; Figure 4A-C). Fluorescence at 24-48 hrs was focal and dispersed throughout the culture (Figure 4A). Using flow cytometry, we found a significant increase in the fluorescence intensity in PHLE cells expressing CX3CR1, compared to CX3CR1 negative cells following RSV infection (Figure 3C). Importantly, pre-incubation of PHLE cells with anti-CX3CR1 antibody prior to RSV infection significantly decreased RSV infection as defined by the number of fluorescence plaques detected (Figure 4A,B). We also found a dose-dependent reduction in the levels of viral transcript production following treatment of PHLE with anti-CX3CR1 antibody (Figure 4C). To confirm the involvement of CX3CR1 in PHLE cell infection by RSV, we incubated the virus with recombinant CX3CR1 (rCX3CR1) prior to infection. We found a significant reduction in RSV-M transcript after preincubation with rCX3CR1 (Figure 4D). Taken together, these results support that CX3CR1 is expressed in pediatric lung epithelial cells, and serves a functional role during infection by RSV. Furthermore, this suggests that CX3CR1 expressing cells are highly susceptible to RSV infection.

**Figure 4.**
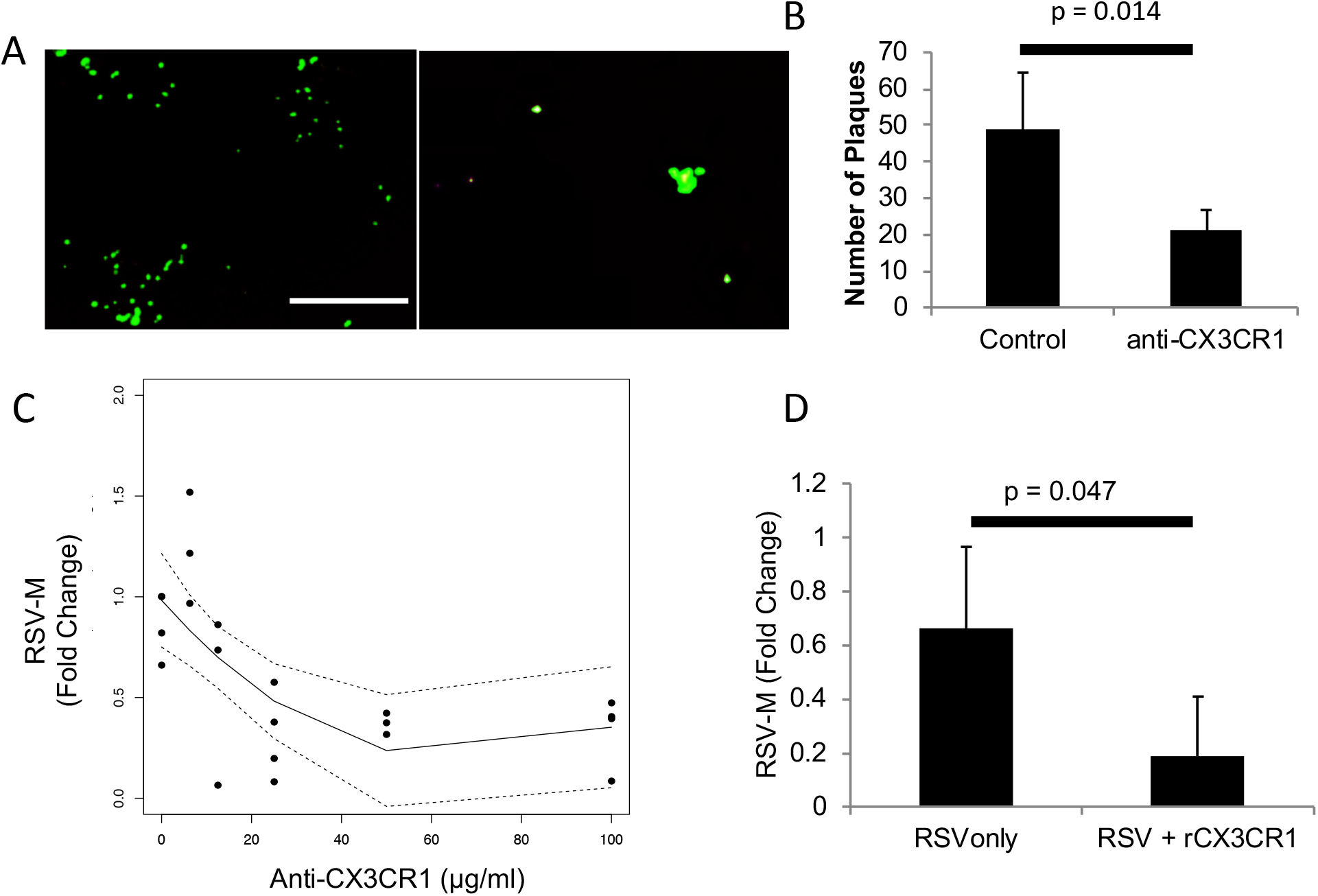
Effect of Blocking CX3CR1 on RSV Infection. (A) Fluorescent images of 24hr post infection PHLE cells infected with GFP expressing RSV virus after cells were incubated for 30 minutes with (right) control antibody or (left) anti-CX3CR1 antibody. (B) RSV plaques detected 24hr post infection with or without CX3CR1 blocking antibody (25ug/ml). (C) Dose-response curve of RSV quantification by qPCR of PHLE cells infected with RSV after preincubation with varying levels of anti-CX3CR1 antibody (n=4). (D) RSV M-protein transcript levels 24hr post infection with or without pre-incubation of RSV with recombinant CX3CR1 protein.

## Discussion

In this report, we show that CX3CR1 mRNA is expressed in airway epithelial cells from the pediatric respiratory tract, including nasal epithelial cells from 53 asymptomatic healthy 1-month old infants and multiple cell types from 11 pediatric lung tissues. Expression appears to be more common in the upper airway than lower airway, but is not always detectable in either location. In vivo, we present evidence that CX3CR1 expression occurs in cells that appear to be ciliated in agreement with other reports using adult cells^18^, in addition to other cells not displaying ciliated cell markers. Using a lung epithelial cell *in vitro* model derived from primary pediatric epithelial cells, we demonstrated that RSV viral loads are highest in cells expressing CX3CR1, and that blocking CX3CR1-RSV interaction significantly reduces levels of RSV infection. Taken together, our results support the expression of CX3CR1 in the airways of children, where it can contribute to productive RSV infection in the airways.

Perhaps, the most surprising finding of our study is the low, and often undetected, RNA expression of CX3CR1 transcript in pediatric lung epithelial cells. Although expression is readily found in other cells in the lung, such as immune cells, epithelial cells appear to express lower levels of CX3CR1 mRNA. Despite this, CX3CR1 protein was detected in the lung epithelium of all subjects we examined. These results are consistent with other reports showing expression of CX3CR1 in normal human bronchial epithelial cell models^6,7^.

We used a novel, physiological model of pediatric lung epithelium. We found that CX3CR1 RNA and protein was expressed in our pediatric model of lung epithelium. The higher levels of CX3CR1 in PHLE cells compared to commonly used cell line, Hep-2, demonstrates the importance of using the correct model to study RSV infection. Upon infection, RSV transcript levels was greatest in PHLE cells expressing CX3CR1. Although we did not discriminate primary vs secondary infection in our model, the increase in RSV transcript levels in CX3CR1 expressing cells supports that RSV preferentially targets CX3CR1 expressing cells. Other studies using adult epithelial cells have demonstrated similar findings^6,7^.

Our results do not demonstrate CX3CR1 is necessary for RSV infection, but contributes to maximal productive infection. Even with high concentrations of blocking antibody, RSV infection was still established. This is consistent with reports showing RSV viruses lacking the G protein can still replicate in vitro, although it is worth noting that these viruses do not cause disease in mice^19^. Additionally, publicly available results from the Lung Molecular Atlas Program profiling project (LungMAP) show expression of heparan sulfate containing proteins (Syndecans) and Nucleolin in lung epithelium^15^. Although expression of heparan sulfate containing proteins in the airways is still debated, our results suggest the presence of at least two receptors capable of mediating productive RSV infections. Given the known binding of RSV to heparin, heparan sulfate, and chondroitin sulfate containing proteins^1^ as well as Nucleolin^2^, future studies are needed to further examine their role in RSV infection of pediatric human lung epithelium.

It should not be overlooked that expression of CX3CR1 in the lung was not restricted to epithelial cells, but rather that many cell types express CX3CR1 (Figure 1C). We found high RNA expression levels of CX3CR1 in immune cells derived from the lung. Given the presence of alveolar macrophages^20^ and T cells^21^ on the apical surface of lung epithelium and the ability of RSV to infect a multiplicity of cells, including CX3CR1 expressing immune cells^22^, it is possible that immune cells play a role in RSV replication^23,24^. Additionally, severe cases of RSV have demonstrated increases of immune cells in the lung^25^. Taken together, further studies are needed to understand the role of CX3CR1 expression on lung infiltrating immune cells and what role this plays in RSV infection and disease severity.

Although there is now considerable evidence that CX3CR1 plays a role as receptor in RSV infection, RSV infection of lung epithelium is more complex then modeled here. Animal models are needed to fully understand the consequence of CX3CR1 expression in cells in the lung. Additionally, our assay did not directly measure attachment of RSV to the lung epithelium via CX3CR1, therefore future studies will be needed to determine the specific role CX3CR1 plays in the airways. Furthermore, transcript levels were low and often undetectable in the epithelial cells of the lung, yet we suggest that sufficient levels of CX3CR1 protein exist despite the low transcript levels. There are no data to suggest that differences in CX3CR1 expression among individuals contributes to the likelihood or severity of infection, but this should be assessed more thoroughly. Overall, our work suggests the CX3CR1 as a therapeutic target, and supports studies aimed at blocking G-protein/host-epithelial interaction as a means to limit RSV infection in children.

## Data Availability

All data is included within the manuscript.

## Acknowledgements

Funding for this work was supported by the University of Rochester Respiratory Pathogen Research Center NIH/NIAID, HHSN272201200005C, the LungMAP Consortium including 1U01HL122700 and the University of Rochester Pulmonary training grant T32-HL066988. The GFP-expressing RSV was kindly provided as a gift from Edward E. Walsh. Human lung tissue was obtained through the non-profit United Network for Organ Sharing. Consent was given by donor or next of kin for the use of tissue in research. The study is approved and overseen by the University of Rochester Research Subjects Review Board (Protocol RSRB00047606), although the work is determined to be non-human research due to tissue donor demise. All material is de-identified to investigators. We greatly appreciate the Donor tissue, precious grifts generously given, supplied through the US DHHS Donation and Transplantation Network and the organizations that link donor families to the scientific community.

